# SARS-CoV-2 seroprevalence and longitudinal antibody response following natural infection in pregnancy: a prospective cohort study

**DOI:** 10.1101/2022.12.28.22284017

**Authors:** Alison L. Drake, Jaclyn N. Escudero, Morgan C. Aurelio, Sascha R. Ellington, Lauren B. Zapata, Romeo R. Galang, Margaret C. Snead, Krissy Yamamoto, Carol Salerno, Barbra A. Richardson, Alexander L. Greninger, Alisa B. Kachikis, Janet A. Englund, Sylvia M. LaCourse

## Abstract

**Importance:** Antenatal care provides unique opportunities to assess SARS-CoV-2 seroprevalence and antibody response duration after natural infection detected during pregnancy; transplacental antibody transfer may inform peripartum and neonatal protection.

**Objective:** Estimate seroprevalence and durability of antibodies from natural infection (anti-nucleocapsid (anti-N) IgG) among pregnant people, and evaluate transplacental transfer efficiency.

**Design:** Seroprevalence study: cross-sectional SARS-CoV-2 antibody screening among pregnant people December 9, 2020-June 19, 2021. Cohort study: Pregnant people screened anti-N IgG+ by Abbott Architect chemiluminescent immunoassay in seroprevalence study or identified through medical records with RT-PCR+ or antigen positive results enrolled in a prospective cohort December 9, 2020-June 30, 2022 to longitudinally measure anti-N IgG responses. We collected cord blood and assessed transplacental transfer of maternally-derived anti-N antibodies.

**Setting:** Three hospitals and 14 affiliated clinics providing antenatal and delivery care, Seattle, Washington metropolitan area.

**Participants:** Seroprevalence study: pregnant people were screened for SAR-CoV-2 anti-N IgG during routine care. Cohort study: Pregnant people with evidence of prior SARS-CoV-2 infection (screened anti-N IgG+ from seroprevalence study or identified with a RT-PCR+ or antigen positive result from medical records) were enrolled in a cohort study to longitudinally measure anti-N IgG responses.

**Exposure(s) (for observational studies):** COVID-19 diagnosis, symptoms, and disease severity.

**Main Outcome(s) and Measure(s):** Presence and durability of SARS-CoV-2 anti-N IgG, transplacental transfer of maternally-derived anti-N IgG.

**Results:** Of 1289 pregnant people screened in the seroprevalence study, 5% (65) tested SARS-CoV-2 anti-N IgG+, including 39 (60%) without prior RT-PCR+ or antigen positive results and 53 (82%) without symptoms. Among 89 participants enrolled in the cohort study, 73 (82%) had anti-N IgG+ results during pregnancy. Among 49 participants with delivery samples 33 (67%) were anti-N IgG negative by delivery. Of 24 remaining anti-N IgG+ at delivery with paired cord blood samples, 12 (50%) had efficient transplacental anti-N IgG antibody transfer. Median time from first anti-N IgG to below positive antibody threshold was 17 weeks and did not differ by prior RT-PCR+ or antigen positive status.

**Conclusions and Relevance:** Maternally-derived SARS-CoV-2 antibodies to natural infection may wane before delivery. Vaccines are recommended for pregnant persons to reduce severe illness and confer protection to infants.

## Introduction

Antenatal care offers a unique opportunity to assess SARS-CoV-2 seroprevalence among pregnant people, including those with previously unknown infection [1]. Prior SARS-CoV-2 seroprevalence studies among pregnant people have been primarily cross-sectional, often focused at delivery in the hospital, and have potentially missed pregnant people infected earlier in pregnancy whose antibody response has waned by the time of delivery, including those with asymptomatic infection or mild disease [2-4].

Natural infection with SARS-CoV-2 during pregnancy may provide some protection against infection during the peripartum period in pregnant people and their infants, but longitudinal immunological responses across the pregnancy-postpartum continuum have not been well characterized. COVID-19 vaccines are recommended for people who are pregnant, recently pregnant (including those who are lactating), trying to become pregnant, or who might become pregnant in the future, regardless of prior infection status [5,6]. Additionally, while COVID-19 vaccines are approved in the United States for children as young as 6 months of age, they are not currently available or being studied in infants aged <6 months [7]. Prospective data on antibody responses following infection during pregnancy and evaluation of transplacental transfer of antibodies to neonates has the potential to provide important information on the durability and duration of maternal and neonatal immunity following natural infection during pregnancy [8].

We conducted a SARS-CoV-2 seroprevalence study among pregnant people in the Seattle, Washington metropolitan area, examined longitudinal SARS-CoV-2 anti-nucleocapsid (anti-N) IgG antibody responses of participants with evidence of natural infection, and measured transplacental transfer of maternally-derived anti-N antibodies. We hypothesized that 5% of pregnant women would have SARS-CoV-2 infection based on anti-N IgG antibody levels and models of SARS-CoV-2 prevalence in the region [9] and the presence of anti-N IgG antibody will be durable through 4 months post-infection based on prior respiratory syncytial virus studies conducted in pregnancy [10-12].

## Materials and methods

### Study setting and participants

#### Seroprevalence study

Pregnant people aged ≥18 years seeking antenatal care at 14 affiliated clinics, or admitted to three labor and delivery units, at University of Washington (UW)-affiliated medical centers were eligible for participation in the seroprevalence study (S1 Table). Healthcare providers obtained informed consent to screen blood samples collected from pregnant people receiving antenatal care during December 9, 2020 - June 19, 2021 for anti-N IgG antibodies to SARS-CoV-2. Healthcare providers offered consent to pregnant people whenever possible; however, we were only able to offer consent to 29% of all people seeking care due to provider inability to offer consent to people who did not access care during the screening period and individual providers’ inability to incorporate consent into clinical care. Samples for screening were derived from samples collected for routine clinical care during December 9, 2020 - June 30, 2021. Individuals who did not have a blood sample available for antibody testing but provided consent for screening (929/2218, 42%) were not screened in the seroprevalence study. History of positive SARS-CoV-2 PCR results and COVID-19 disease severity and symptoms were abstracted from the electronic medical record for people who screened positive for anti-N IgG.

**S1 Table. Study Sites**

#### Prospective cohort study

Pregnant people with evidence of prior SARS-CoV-2 infection (with either documented RT-PCR positive results via medical record reviews during pregnancy or within 6 months before pregnancy or with anti-N IgG positive results from the seroprevalence study) during December 9, 2020 - June 8, 2022 were eligible to enroll in a cohort study evaluating longitudinal SARS-CoV-2 anti-N IgG responses. Beginning in January 2022, pregnant people who self-reported a positive rapid antigen test for SARS-CoV-2 infection were also eligible to enroll. Participants identified with a RT-PCR or rapid antigen positive test had blood samples for the prospective cohort collected following informed consent. For participants identified through the seroprevalence study, the seroprevalence result served as the enrollment sample. All participants were scheduled for follow-up blood sample collection at 1, 2, 3 months post-enrollment and delivery (including maternal and cord blood). Additional blood samples were collected at 1-2, 2-4, and 6 months postpartum if sample collection dates did not fall within the post-enrollment sample collection windows. All blood samples were tested for SARS-CoV-2 anti-N IgG antibodies. COVID-19 disease severity and symptoms were abstracted from the electronic medical record and classified as asymptomatic, mild, severe, or critical; disease severity was reported for the initial infection for individuals who were known to have multiple SARS-CoV-2 infections [13].

### Laboratory methods

#### SARS-CoV-2 anti-N IgG serology

Samples collected for SARS-CoV-2 serology were tested using the Abbott Architect chemiluminescent immunoassay (CMIA), an automated qualitative test designed to detect anti-N IgG to SARS-CoV-2 (Abbott, Abbott Park, Illinois, USA) at UW. This assay has high sensitivity (100%, 17 days post-infection) and specificity (>99.9%) in the two months after SARS-CoV-2 infection [14-16]. Samples with an Abbott index ≥1.4 were considered positive per manufacturer recommendations [17].

#### Exposure and outcome variables

The primary exposures were SARS-CoV-2 diagnosis, symptoms, and disease severity; the primary outcomes were presence and durability of SARS-CoV-2 anti-N IgG, and transplacental transfer of maternally-derived anti-N IgG antibodies.

#### Statistical analysis

Transplacental transfer ratios were calculated as the Abbott index from cord blood collected at delivery divided by the Abbott index from maternal blood collected at delivery with a ratio of ≥1 considered as efficient transplacental transfer. Vaccination status based on medical record abstraction and/or self-report was classified as follows: partial with one dose of an mRNA vaccine, fully vaccinated if two doses of an mRNA vaccine or one dose of a viral vector vaccine, and boosted if three doses of an mRNA vaccine (or at least one dose plus a viral vector vaccine) or two doses of viral vector vaccine. Wilcoxon rank sum tests were used to compare distributions of continuous variables. Prevalence ratios were calculated to assess potential sources of bias in people who consented versus declined and those who were screened vs did not have a blood sample collected in the seroprevalence study. We assessed co-factors of SARS-CoV-2 anti-N IgG positive results which were calculated using generalized linear models (GLMs) with Poisson family and log link. Kaplan-Meier analysis was used to calculate time to anti-N IgG Abbott below the positive threshold (index <1.4); differences in curves by PCR/antigen status were assessed using the log-rank test. Cox proportional hazards regression models were used to explore potential covariates of time to anti-N IgG below the threshold. Anti-N IgG Abbott index results were log_10_ transformed to reduce skewness, and general estimating equations (GEE) with a Gaussian link and robust standard errors were constructed to measure the rate of change in log_10_ anti-N IgG response over time since first anti-N IgG positive result and to assess potential co-factors. Potential co-factors for the Cox and GEE models included: trimester of infection, an interaction between pregnancy status and time since first anti-N IgG positive result, presence of symptoms, disease severity, and vaccination status.

#### Sample size

Seroprevalence study: With a sample size of 1268 pregnant people, we have 1.22% precision to detect seroprevalence based on anti-N IgG results of 5% (95% CI 3-7%) by the end of pregnancy (i.e., at the time of delivery admission); an additional 21 people were enrolled by healthcare providers after the target sample size was reached; therefore, the overall sample size was 1289.

Prospective cohort study: We calculated a sample size of 50 pregnant people with evidence of SARS-CoV-2 would be required to measure correlates of infection during pregnancy (to be presented in a future manuscript) with effect sizes of 2.5-3.4 for a range of co-factors for infection with prevalence ranging 10-40%, assuming 80% power, alpha=0.05, and two-sided testing; however, we continued to accrue additional infections in the cohort due to the convenience sample of pregnant people with virologic evidence of SARS-CoV-2 infection available during the study time frame and reported to the study by healthcare providers.

#### Ethics statement

This study was approved by UW Institutional Review Board and UW Medicine Valley Medical Center Research Oversight Committee. The activity was reviewed by the US Centers for Disease Control and Prevention (CDC) and was conducted consistent with applicable federal law and CDC policy. All participants provided written informed consent prior to study participation.

## Results

### SARS-CoV-2 seroprevalence study

Overall, we identified 8632 pregnant people who received antenatal care or delivered at UW enrollment sites during the seroprevalence study enrollment period (Fig 1, S1 Table); 2690 pregnant people (31%) were offered participation and consent for the study (2218 consented and 472 declined). Among the 2218 who consented to blood screening for SARS-CoV-2 anti-N IgG, 1289 (58%) had blood samples tested. There were some significant differences in pregnant people who consented to screening and had samples tested by age, race, and ethnicity (p<0.05, S2 Table); notably Black race was more frequently reported among those who declined screening (19%) than those who consented to screening (7%). Among those who consented, Black race was less frequently reported among those who did not have blood tested (1%) than those who were screened (8%). Among pregnant people screened for anti-N IgG, the median age was 32 years (interquartile range [IQR] 29-36) with a median gestational age at screening of 16 weeks (IQR 11-38) Table 1.

**Table 1.** Baseline characteristics of pregnant people screened for SARS-CoV-2 anti-N IgG, by anti-N IgG result.

|  | Screened for anti-N IgG (n=1289) | Anti-N IgG- (n=1224) | Anti-N IgG+ (n= 65) | PR<sup>c</sup> (95% CI) | p-value |
| --- | --- | --- | --- | --- | --- |
|  | *n (%) or median (IQR)* |  |  |  |  |
| Age<sup>a</sup> | 32 (29-36) | 32 (29-36) | 29 (26-34) | 0.92 (0.87-0.97) | 0.001 |
| Gestational age (weeks) at screening<sup>b</sup> | 16 (11-38) | 15 (11-38) | 18 (11-38) | 1.00 (0.99-1.02) | 0.74 |
| Race |  |  |  |  |  |
| American Indian/Alaska Native | 17 (1) | 15 (1) | 2 (3) | 3.99 (1.02-15.70) | 0.047 |
| Asian | 256 (20) | 252 (21) | 4 (6) | 0.53 (0.18-1.53) | 0.24 |
| Black | 103 (8) | 90 (7) | 13 (20) | 4.25 (2.20-8.22) | <0.0001 |
| Native Hawaiian/Pacific Islander | 31 (2) | 26 (2) | 5 (8) | 5.48 (2.21-13.56) | <0.0001 |
| White | 713 (55) | 692 (57) | 21 (32) | REF |  |
| Other | 112 (9) | 94 (8) | 18 (28) | 5.46 (3.00-9.92) | <0.0001 |
| Not reported | 57 (4) | 55 (4) | 2 (3) | - | - |
| Hispanic ethnicity | 138/1244 (11) | 120/1244 (10) | 18/64 (28) | 3.13 (1.87-5.25) | <0.0001 |

PR, prevalence ratio comparing anti-N IgG+ vs anti-N IgG-. CI, confidence interval. Anti-N IgG tested using Abbott Architect chemiluminescent immunoassay (CMIA), a semi-qualitative test designed to detect anti-nucleocapsid (anti-N) IgG of SARS-CoV-2. IgG+ if Abbott index $\geq 1.4$ .
<sup>a</sup>Missing age for 9 IgG- people.
<sup>b</sup>Missing gestational age for 9 people (8 IgG- and 1 IgG+ people).
<sup>c</sup>per 1 unit increase

**Fig 1.**
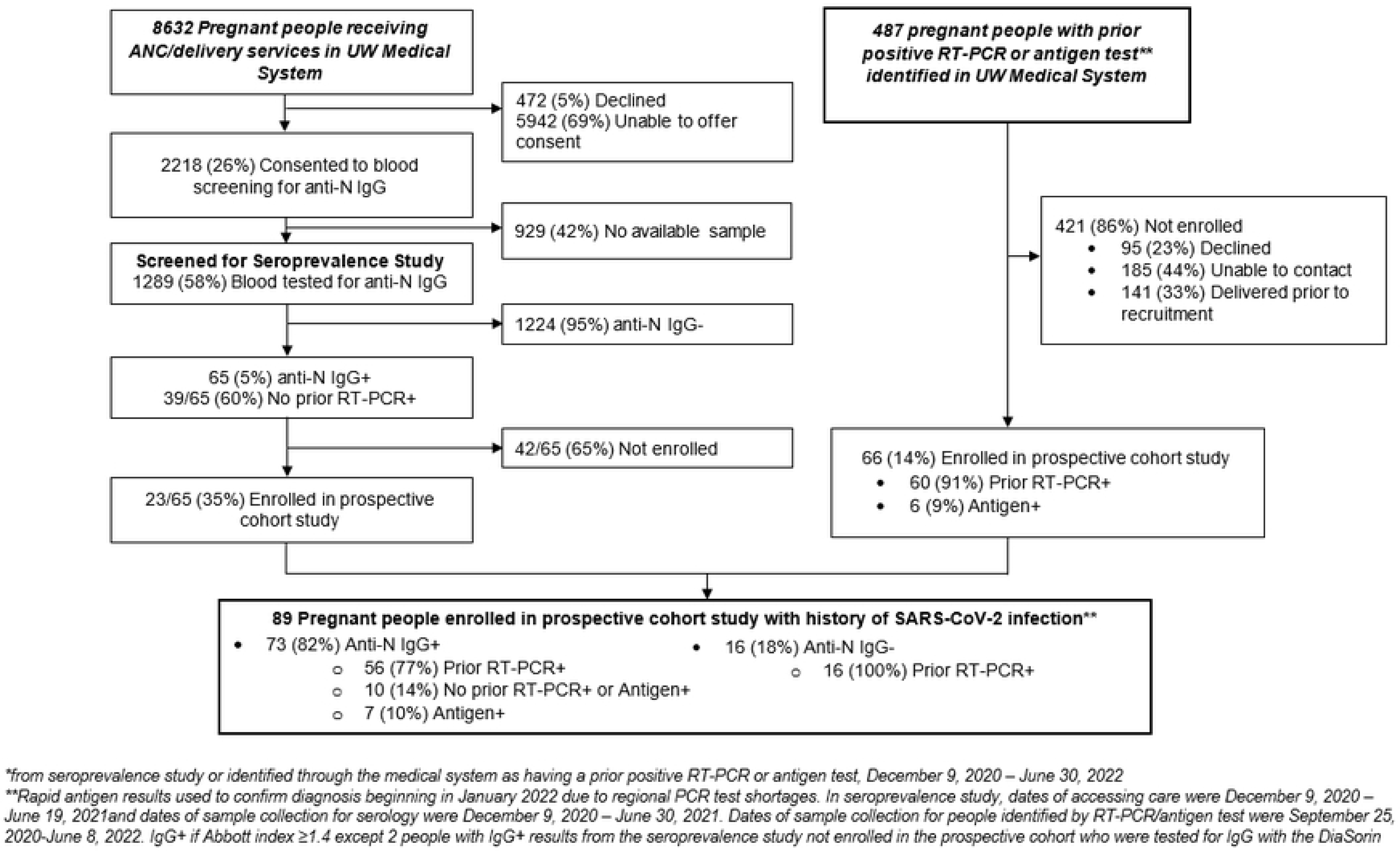
Flowchart of pregnant people enrolled in prospective cohort study with a history of SARS-CoV-2 infection*.

**S2 Table. Pregnant people who consented to screening and had samples tested by age, race, and ethnicity**

SARS-CoV-2 seroprevalence among pregnant people screened was 5% (65/1289), of whom 60% (n=39) did not have a prior RT-PCR positive test result during pregnancy documented in their medical record. Monthly seroprevalence was significantly different over time (p=0.03), peaking in December 2020 at 8.2% prior to the World Health Organization variants of concern becoming dominant in the area, and was lowest in April 2021 before the Delta variant became dominant (S1A,B Fig) [18]. There were significant differences in seroprevalence by race and ethnicity Table 1. Among those who tested negative for anti-N IgG, 57% were White compared with 32% of those testing positive for anti-N IgG. Compared to White pregnant people, seroprevalence was at least 3 times as high among those who were Black, Native Hawaiian or Pacific Islander, American Indian or Alaska Native, or identified as another race (p<0.05 for all). Seroprevalence was also 3 times as high among pregnant people of Hispanic versus non-Hispanic ethnicity. Older pregnant people were less likely to test anti-N IgG positive with each year increase in age associated with 8% lower seroprevalence (PR: 0.92, 95% CI for PR: 0.87-0.97). Among 65 pregnant people identified in the seroprevalence study with anti-N IgG positive results, 23 (35%) had symptoms (21 mild, 2 severe), 2 (3%) of whom were hospitalized for COVID-19. RT-PCR dates were available for 22 of 26 pregnant people with a RT-PCR positive result, with a median time between RT-PCR positive date and blood collection date for anti-N IgG of 7 weeks (IQR 4-15); 4 people were RT-PCR positive before pregnancy.

**S1A Fig. SARS-CoV-2 anti-N IgG screening status of pregnant people participating in the seroprevalence study**

**S1B Fig. Anti-N IgG seroprevalence by month**

Median Abbott index among 65 pregnant people with anti-N IgG positive results (Abbott index ≥ 1.4) and available Abbott index in the seroprevalence study was 3.18 (IQR 2.06-5.00). Median Abbott index was significantly higher among those with COVID-19 symptoms reported in the medical record than without (median 4.39, IQR 3.18-5.42 versus median 2.49, IQR 1.92-4.30; respectively; p=0.02) and among those with RT-PCR positive results than without (median: 4.19, IQR 2.87-5.42 versus 2.49, IQR 1.89-4.16; respectively; p=0.02). Fifteen participants received a COVID-19 vaccine (14 fully vaccinated, 1 partially vaccinated) prior to initial anti-N IgG positive test, but no differences in anti-N IgG Abbott index were detected between people who were and were not vaccinated (data not shown).

### Prospective cohort study

We enrolled 89 pregnant people with evidence of prior SARS-CoV-2 in the prospective cohort study; 23 (26%) from the seroprevalence study and 66 (74%) identified through medical records (Fig 1). Median age was 32 years (IQR 30-35) and median gestational age at first blood sample collection was 25 weeks (IQR 16-36). Less than 5% of participants identified as either non-binary or did not report their gender; all other participants identified as women. Overall, 16 (18%) were anti-N IgG negative, all with prior RT-PCR positive results. Among 73 of 89 (82%) people with anti-N IgG positive results, most (n=56, 77%) also had a prior RT-PCR positive result, 10 (14%) positive by anti-N IgG alone, and 7 (10%) also had a prior positive antigen test but no prior RT-PCR positive result.

### Pregnant people with SARS-CoV-2 anti-N IgG positive results

Of 89 pregnant people enrolled in the prospective cohort, 73 (83%) had anti-N IgG positive results documented at any time during the study Table 2. Among 68 people who were pregnant when the first anti-N IgG positive result was collected, the median gestational age was 22 weeks (IQR 15-36). Most (n=56, 77%) had a prior positive RT-PCR result (5 recorded prior to pregnancy [median 8 weeks before; range 0.6-24 weeks], data not shown), 55 (75%) reported having symptoms, and 11 (15%) had neither reported symptoms or a prior RT-PCR positive result (data not shown). Among the 55 people with symptoms reported, 71% (n=55) had mild symptoms and 4% (n=3) had severe symptoms; 3 were hospitalized for COVID-19. Median Abbott index at the first anti-N IgG positive sample was 3.53 (IQR 2.33-5.12, range [data not shown] 1.43–9.87), and was not significantly different between those with (3.72, IQR 2.53-5.12) and without (2.65, IQR 1.97-5.00) prior RT-PCR positive results (p=0.34) (data not shown). Median Abbott index at the first anti-N IgG positive sample was also not significantly different between those who were symptomatic versus asymptomatic (3.73 vs 2.5, p= 0.19) (data not shown). Among 57 pregnant people with known dates for their prior RT-PCR positive result, the median time from RT-PCR positive result to first available anti-N IgG positive result was 6 weeks (IQR 4-12) (data not shown).

**Table 2.** Baseline characteristics of pregnant people enrolled with prior SARS-CoV-2 infection and anti-N IgG+.

| | Anti-N IgG+ (N=73) | Anti-N IgG+ with $\geq 1$ follow-up anti-N IgG result (N=61) |
| --- | --- | --- |
|  | n (%) or Median (IQR) |  |
| Age (years) | 32 (29-34) | 32 (30-35) |
| Gestational age at first blood collection (weeks)<sup>a</sup> | 22 (15-36) | 20 (14-33) |
| Prior RT-PCR+ result | 56 (77) | 44 (72) |
| Abbott Index at 1<sup>st</sup> anti-N IgG+<sup>b</sup> | 3.53 (2.33-5.12) | 3.53 (2.35-4.89) |
| COVID-19 severity |  |  |
| Asymptomatic | 18 (25) | 18 (30) |
| Mild | 52 (71) | 42 (69) |
| Critical | 0 (0) | 0 (0) |
| Severe | 3 (4) | 1 (2) |
| Hospitalized for COVID-19 | 3 (4) | 1 (2) |
| Vaccination status at 1<sup>st</sup> anti-N IgG+ <sup>c</sup> |  |  |
| None | 43 (59) | 36 (59) |
| Partial | 3 (4) | 3 (5) |
| Fully | 17 (23) | 13 (21) |
| Boosted | 10 (14) | 9 (15) |

<sup>a</sup>5 people (1 with $\geq 1$ follow-up) whose first blood collection was after delivery <sup>b</sup>Abbott index at 1<sup>st</sup> anti-N IgG+ $\geq 1.4$ . <sup>c</sup>Vaccination status was classified as partial with one dose of an mRNA vaccine or missing vaccine type; fully vaccinated if two doses of an mRNA vaccine, two doses with missing vaccine type, or one dose of a viral vector vaccine; and boosted if three vaccine doses of any type (three mRNA vaccines, two doses plus a viral vector vaccine, or three doses with one or missing vaccine types) or two doses of viral vector vaccine.

Among 61 pregnant people with an anti-N IgG positive result and at least one subsequent blood sample, the median time from the first positive anti-N IgG result to anti-N IgG below the positive threshold was 17 weeks (95% CI 12-27) and was similar regardless of whether they had a prior RT-PCR positive result (median 17 weeks for both) (Fig 2a). Median time from first RT-PCR positive result to anti-N IgG below the positive threshold was 28 weeks (95% CI 19-33) (Fig 2b). The average rate of change in log_10_ Abbott index per month was: 0.02 increase since first blood collection (Fig 3a), 0.02 decrease since RT-PCR positive result (Fig 3b), and 0.02 decrease since RT-PCR positive or antigen positive result (Fig 3c). Log_10_ Abbott index was significantly lower at the first blood collection for people who were in the second trimester (p=0.02), and a trend toward lower among people in their third trimester (p=0.09), compared to the first trimester. The average rate of change in log_10_ Abbott index per month was significantly higher, with increases for people in both the second and third trimester compared to at or before the first trimester (p=0.002 and p=0.0001, respectively). There were no differences in the rate of change in log_10_ Abbott index by disease severity/symptoms, pregnant versus postpartum status, or vaccination status (results not shown).

**Fig 2A.**
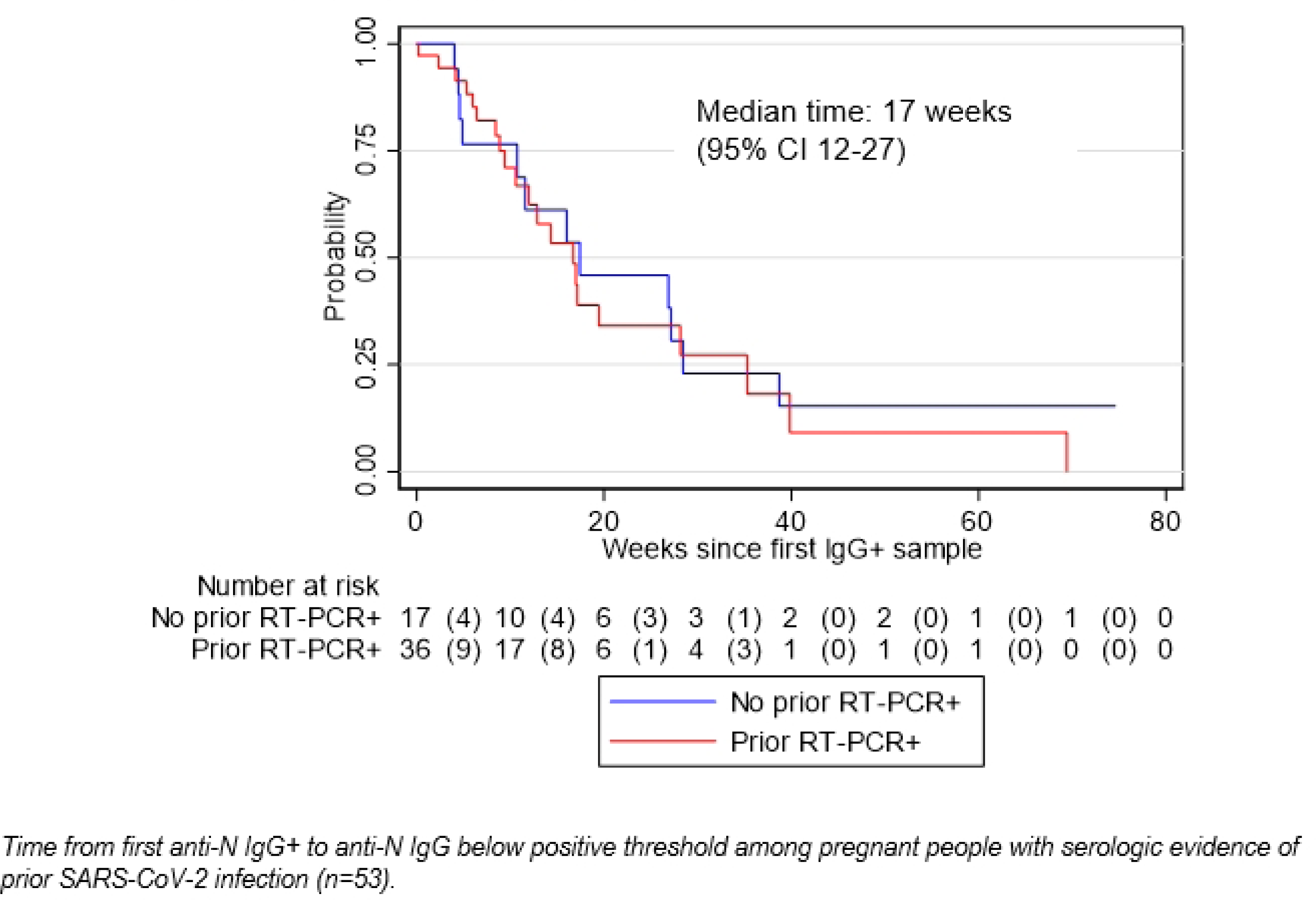
Time from first anti-N IgG+ to anti-N IgG below positive threshold. Anti-N IgG index below positive threshold based on Abbott index <1.4.

**Fig 2B.**
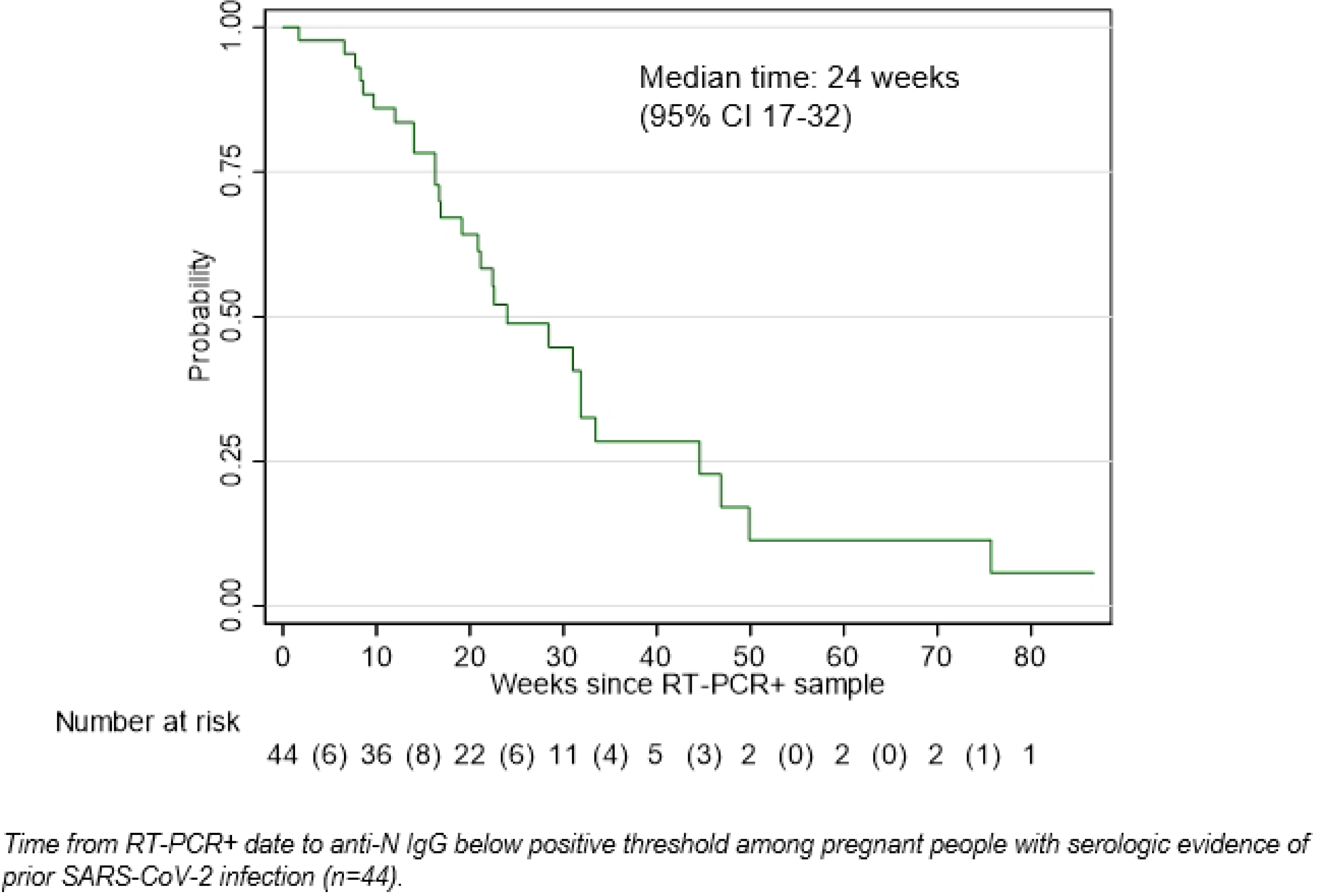
Time from RT-PCR+ to anti-N IgG below positive threshold. Anti-N IgG index below positive threshold based on Abbott index <1.4.

**Fig 3A.**
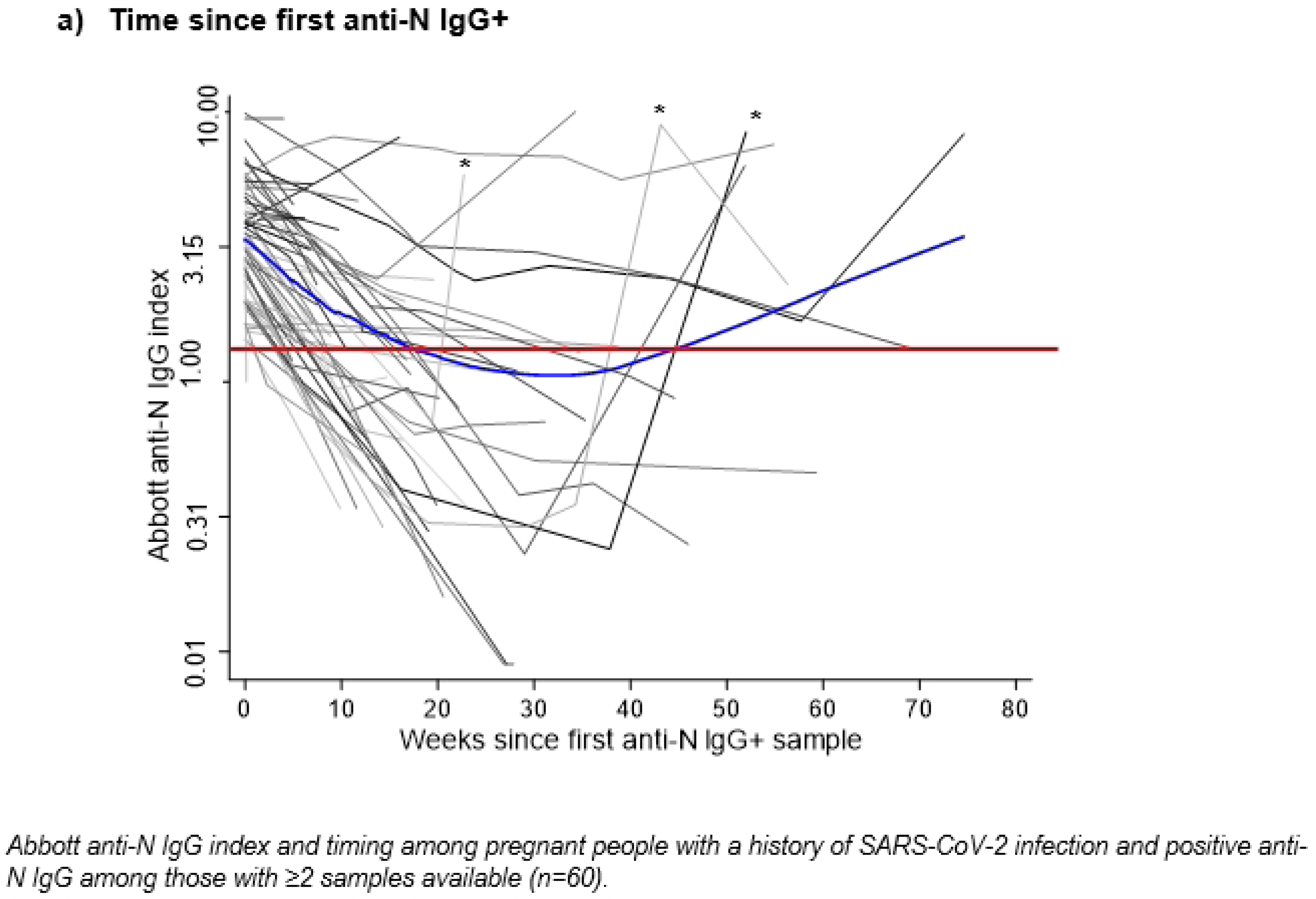
Abbott anti-N IgG index and timing. *First sample collected following reinfection with SARS-CoV-2, confirmed with subsequent RT-PCR+ or antigen positive result (n=3). Red line represents Abbott index for positive threshold (log10 transformed value of 1.4. Blue line represents lowess smoother; excludes samples collected at or after SARS-CoV-2 infection.

**Fig 3B.**
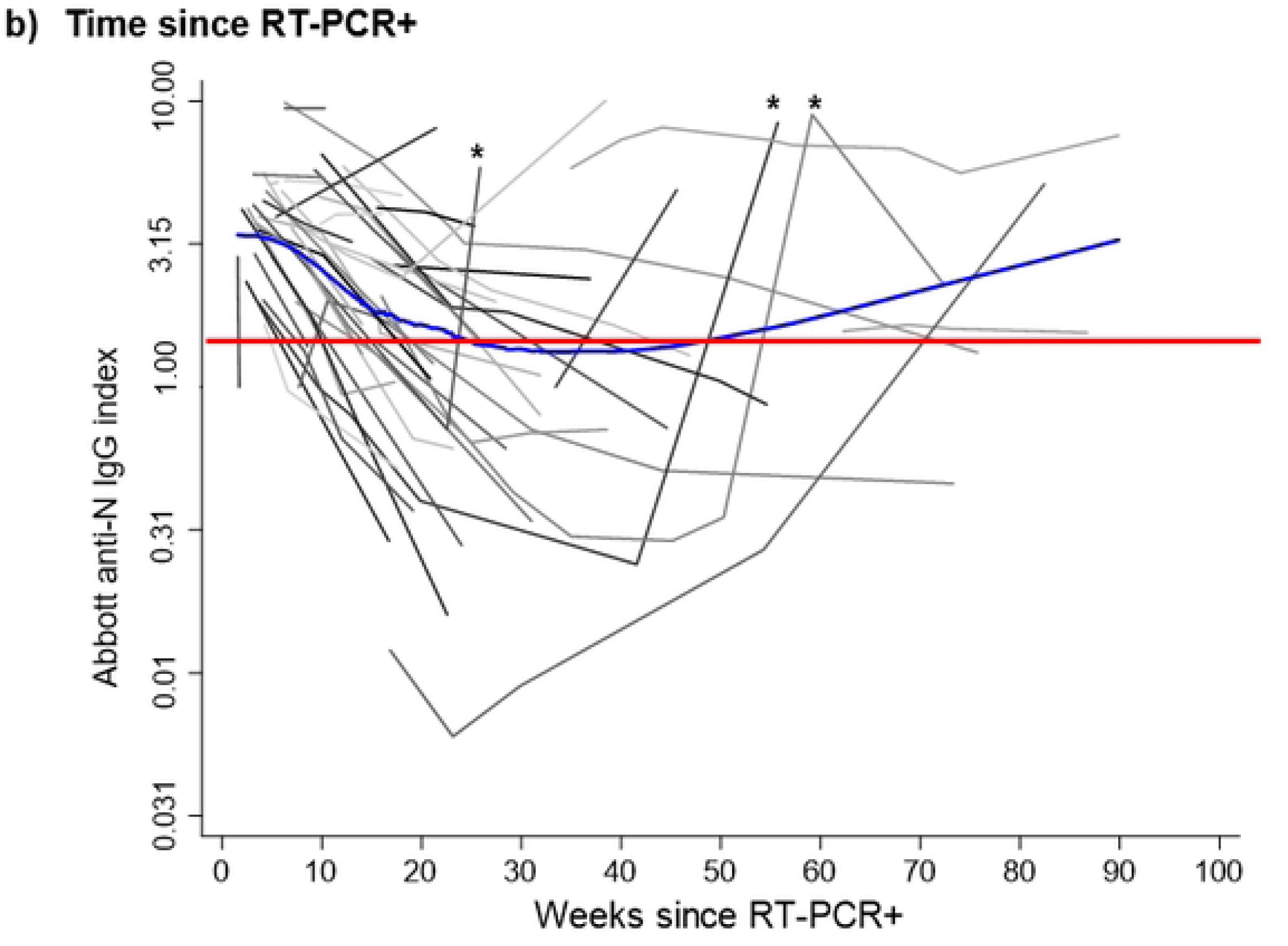
Time since RT-PCR+.

**Fig 3C.**
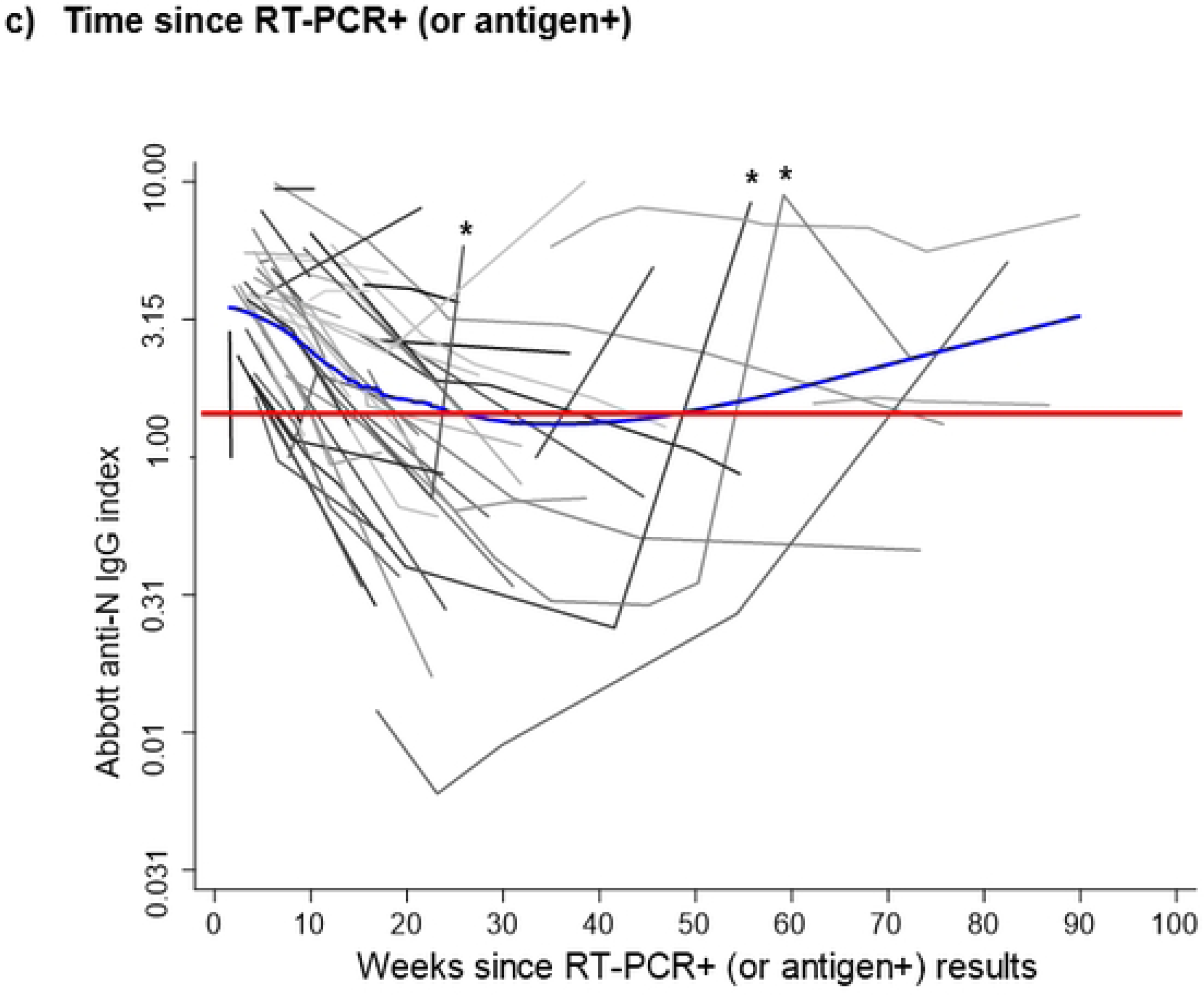
Time since RT-PCR+ (or antigen+).

### Anti-N IgG at delivery and transplacental antibody transfer

Maternal blood samples were collected at delivery from 49 people with prior anti-N IgG positive results; 33 (67%) remained anti-N IgG positive and 16 (33%) were anti-N IgG negative at delivery. Paired maternal and cord blood were collected from 37 of 49 participants at delivery. Delivery samples for people with paired cord blood were collected at a median of 8 weeks after the first anti-N IgG positive result (n=37) and a median 12 weeks after the RT-PCR positive result (n=30). Among 24 participants with anti-N IgG positive results at delivery and available cord blood results, most (n=21, 88%) cord blood samples were also anti-N IgG positive (Fig 4). However, half (n=12, 50%) had evidence of efficient transplacental transfer of anti-N IgG with transfer ratio of ≥1. The corresponding median IgG index for maternal blood and cord blood samples above the positive threshold was 3.66 (IQR 2.25-5.27) and 3.06 (IQR 2.73-4.52), respectively. Among participants remaining anti-N IgG positive at delivery, the median placental transfer ratio of maternally derived anti-N IgG was 0.94 (IQR 0.73-1.31). Maternal samples collected at delivery that were anti-N IgG negative were collected at a median of 19 weeks after the initial anti-N IgG positive result and 19 weeks after the prior RT-PCR positive result.

**Fig 4.**
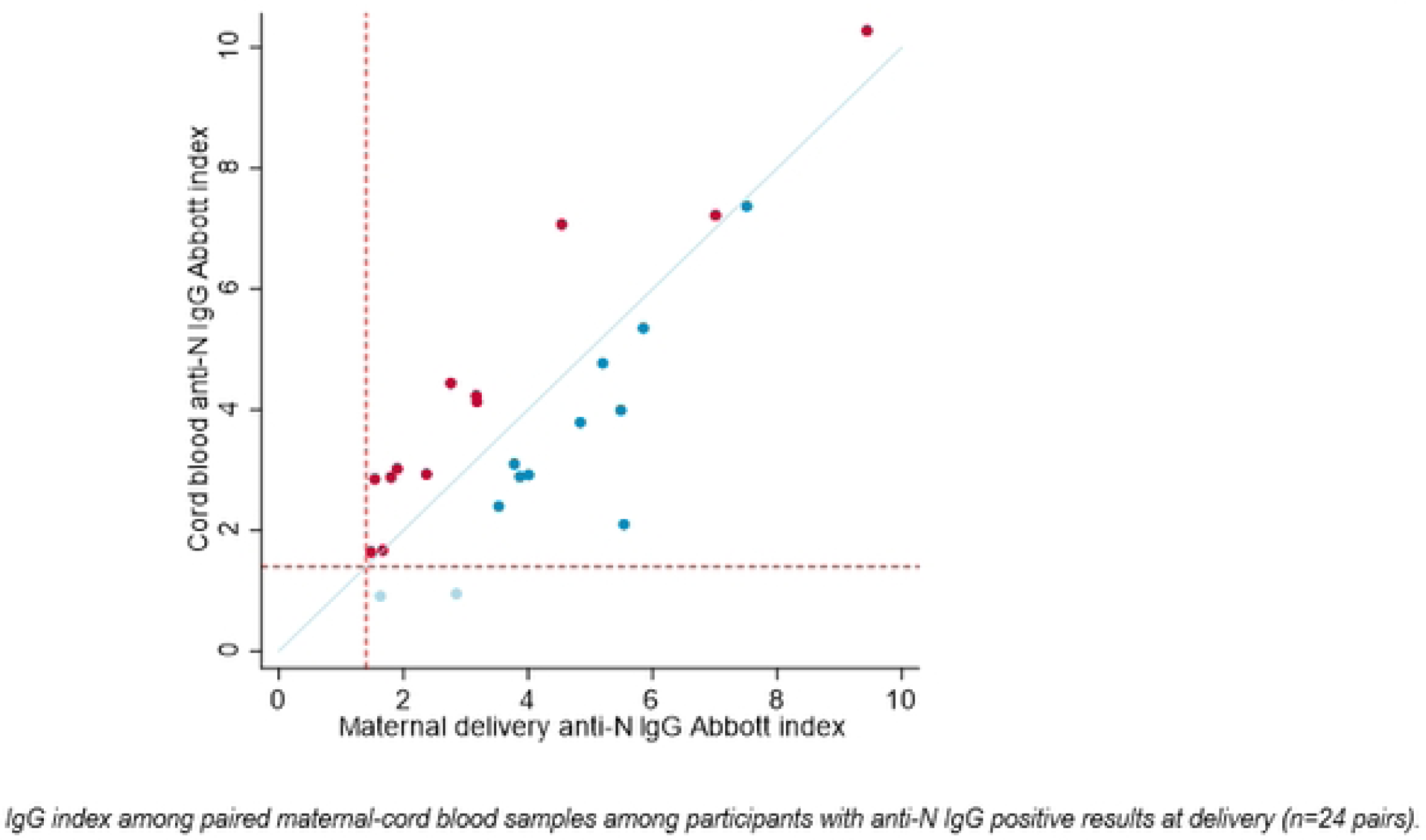
IgG index among paired maternal-cord blood samples. Transplacental transfer ratio (infant anti-N IgG index/maternal delivery anti-N IgG index) is ≥1 for circles on or above the blue line (red), <1 for circles below the blue line (blue); circles below the dashed red lines (threshold for anti-N IgG positive [≥1.4]) are cord blood samples below the anti-N IgG positive threshold (light blue).

## Discussion

SARS-CoV-2 seroprevalence among 1289 pregnant people was 5%, in the Seattle metropolitan area between December 2020 and June 2021, with highest prevalence of ∼8% in December 2020-January 2021. Over half of pregnant people identified with serological evidence of natural SARS-CoV-2 infection did not have a prior RT-PCR or antigen positive test documented in their medical record. Among all people with a prior SARS-CoV-2 infection enrolled in the cohort study, 15% were both asymptomatic and unaware they were previously infected with SARS-CoV-2. While our estimates of asymptomatic (and mild) infections among pregnant people with RT-PCR+ results are similar to other pregnant cohorts [19,20], our assessment of anti-N IgG enabled us to characterize longitudinal antibody responses among people with less severe infections that may be missed in other seroprevalence SARS-CoV-2 studies conducted in pregnancy. In our study, we found that pregnant people with serologic evidence of SARS-CoV-2 were more likely to be Black, Native Hawaiian/Pacific Islander, or American Indian/Alaska Native race or Hispanic ethnicity; these observed disparities concur with results from prior studies of SARS-CoV-2 infection conducted among pregnant people King County, WA (location of Seattle metropolitan area) [21] and in other parts of the U.S. [3,21-23]. We found anti-N IgG responses waned over time following natural infection; these results concur with other studies of waning anti-N IgG levels in non-pregnant cohorts, particularly those using the Abbott Architect platform [24-25]. While two-thirds of pregnant people with anti-N IgG antibodies in pregnancy remained anti-N IgG positive at delivery, with most cord blood samples also remaining anti-N IgG positive, less than half of participants remaining anti-N IgG positive at delivery had evidence of efficient transplacental transfer of maternal anti-N IgG with ratio of cord to maternal anti-N antibody ratio of ≥1. Assuming that pregnant people who were anti-N IgG negative at delivery would have had corresponding negative cord samples, we estimate that only 28% of people who had prior evidence of anti-N IgG antibodies efficiently transferred anti-N IgG transplacentally. These results concur with prior studies demonstrating correlations between maternal IgG and cord blood IgG concentrations [3]. Our findings have important implications for conveying information about potential protection following natural infection during (or before) pregnancy. Natural infection may contribute to decisions to delay or decline vaccination under the assumption that natural infection provides sufficient immunity against maternal reinfection, infant infection, and COVID-19 morbidity and mortality. Median time to a decrease in anti-N IgG below the threshold for positive was 4 months after anti-N IgG was first detected and 6.5 month after testing positive by RT-PCR. While this duration was similar to some studies conducted among non-pregnant cohorts [26] and shorter than others [27], it corresponds to one-third of pregnant people with previous SARS-CoV-2 infection no longer having positive anti-N IgG antibodies by the time of delivery – a critical time (including for late pregnancy) for transplacental transfer of protective antibodies to infants.

Some studies of transplacental transfer of maternal IgG to SARS-CoV-2 suggest efficiency is greatest when infection occurs during the second trimester, while others have found transfer efficiency is higher in the third trimester [28]. Differences in these studies may be attributed to duration of infection prior to delivery, with potential for antibody waning if infection occurs early in pregnancy, and insufficient time to mount a robust immune response if infection occurs too close to delivery. Alternatively, efficiency of antibody transfer may be related to severity of disease, or disease severity and timing of infection. Continued collection of data on potential correlates of infant protection from severe illness by maternally derived antibodies (whether from natural infection or vaccination) will be important to evaluate the degree of protection afforded by maternal immunity.

Our study had several strengths. The seroprevalence study included a large catchment area (medical facilities that capture >6200 deliveries annually) and a similar racial and ethnic distribution to King County and the general Seattle metropolitan area. Since pregnant people are generally healthy, our estimate may be representative of general population prevalence [29]. Characterization of antibody responses over time following natural infection includes pregnant people who may have had undiagnosed, mild and/or asymptomatic infection identified through the seroprevalence study. Similarly, we collected cord blood and measured transplacental antibody transfer of anti-N IgG in a cohort of pregnant people that included infections that were less severe. Finally, we were able to longitudinally characterize maternal infection for at least 6 months, throughout pregnancy and the early postpartum period.

Our study also has some limitations. While the Abbott Architect anti-N IgG sensitivity is high in the weeks to months immediately after infection, the assay preferentially detects low affinity antibodies known to wane [24,30] and it may fail to detect individuals infected with SARS-CoV-2 whose anti-N IgG response waned below the positive threshold prior to sample collection [25]. Even with detectable levels of anti-N IgG, the degree of potential protection offered by anti-N (vs anti-spike or other immune markers) to neonates or young infants is unknown. Estimates of >50% of pregnant people identified as having serologic evidence of natural infection without a known prior infection history of were based on medical record review; it is possible that some participants may have had a prior positive RT-PCR or antigen test that was not documented in their medical record. During later periods of enrollment for the prospective cohort a self-reported positive antigen test was considered evidence of prior infection, which could introduce bias not present earlier in the enrollment period. While we were unable to determine the timing of infection for 10% of women without a prior RT-PCR or antigen positive result, the duration of anti-N IgG response at or above the Abbott index threshold were similar among those with and without prior RT-PCR or antigen positive results, which suggest our results were robust despite this limitation. Pregnant people with serologic evidence of SARS-CoV-2 were captured when different dominant variants were circulating in the area, thus their antibody responses are likely based on a variety of variants; however, we were unable to characterize responses by variant. In addition, following the circulation of Omicron variants in late 2021 and early 2022, the time between identification of RT-PCR+ or antigen positive result and first blood draw was short (3.5 weeks) compared to 5.7 weeks prior to widespread circulation of Omicron. Lastly, differences in age, race, and ethnicity of participants who consented versus declined and those who did and did not have blood samples available for testing may have biased the results; similarly, there may be differences in people who were and were not offered participation in the seroprevalence study resulting in selection bias. We anticipate SARS-CoV-2 seroprevalence may be biased towards the null due to the larger proportion of participants from minority race and ethnic groups who declined blood screening for anti-N IgG, and these groups are more likely to have become infected with SARS-CoV-2 during the study period.

## Conclusions

In conclusion, we found that anti-N IgG levels following natural infection during pregnancy wane over time, with only two-thirds of pregnant people remaining with anti-N IgG positive results at delivery and less than half of these individuals had evidence of efficient transplacental anti-N antibody transfer to their infants. More information is needed on the timing and severity of maternal natural infection provided by anti-N IgG, as well as other immune markers, to better understand the transfer and potential for protection to infants. Anti-N IgG levels may not indicate a sustained immunological response. These data further support the use of vaccines during pregnancy among people, even among those who have previously been infected with SARS-CoV-2 [5,6].

## Data Availability

Yes, it will be made available upon request.

## Acknowledgement

The US Centers for Disease Control and Prevention (CDC) provided technical assistance related to analysis and interpretation of data and writing the report. The findings and conclusions in this report are those of the authors and do not necessarily represent the official position of the US Centers for Disease Control.

